# Automated Localization and Segmentation of Mononuclear Cell Aggregates in Kidney Histological Images Using Deep Learning

**DOI:** 10.1101/19002634

**Authors:** Dmytro S. Lituiev, Sung Jik Cha, Aaron Chin, Benjamin S. Glicksberg, Andrew Bishara, Dejan Dobi, Ruizhe (Ryan) Cheng, Jae Ho Sohn, Zoltan Laszik, Dexter Hadley

## Abstract

Allograft rejection is a major concern in kidney transplantation. Inflammatory processes in patients with kidney allografts involve various patterns of immune cell recruitment and distributions. Lymphoid aggregates (LAs) are commonly observed in patients with kidney allografts and their presence and localization may correlate with severity of acute rejection. Alongside with other markers of inflammation, LAs assessment is currently performed by pathologists manually in a qualitative way, which is both time consuming and far from precise. Here we present the first automated method of identifying LAs and measuring their densities in whole slide images of transplant kidney biopsies. We trained a deep convolutional neural network based on U-Net on 44 core needle kidney biopsy slides, monitoring loss on a validation set (*n*=7 slides). The model was subsequently tested on a hold-out set (*n*=10 slides). We found that the coarse pattern of LAs localization agrees between the annotations and predictions, which is reflected by high correlation between the annotated and predicted fraction of LAs area per slide (Pearson R of 0.9756). Furthermore, the network achieves an auROC of 97.78 ± 0.93% and an IoU score of 69.72 ± 6.24 % per LA-containing slide in the test set. Our study demonstrates that a deep convolutional neural network can accurately identify lymphoid aggregates in digitized histological slides of kidney. This study presents a first automatic DL-based approach for quantifying inflammation marks in allograft kidney, which can greatly improve precision and speed of assessment of allograft kidney biopsies when implemented as a part of computer-aided diagnosis system.

## Introduction

End-to-end deep learning (DL) methods has approached human performance in multiple computer vision tasks ^1,2^ and have more recently been successfully utilized in many biomedical domains^3–5^. Convolutional neural networks (CNN) are a class of artificial neural networks commonly used in DL approaches for image analysis and are composed of multiple layers of convolutional filters and nonlinear activation units to extract meaningful features. These features are then used to produce a classification for each input image. CNNs are especially well equipped for image classification tasks due to the spatial invariance of the learned features and the presence of nonlinear activation units that allow for the learning of complex features^1^. In recent years, deep learning using CNNs has been shown to achieve high performance in image segmentation tasks^6,7^, in which each pixel of an image is assigned a discrete class. By assigning each image pixel a discrete class label, the image can be segmented into distinct regions of interest. The use of CNNs have thus recently gained traction in biomedical segmentation^6^ and digital pathology tasks, such as the detection and localization of breast cancer and its metastases in lymph nodes^8,9^. Based on such successful applications, a number of DL-based approaches have been approved for clinical use in the USA and other countries and are already contributing to higher precision and efficiency in many domains of diagnostics and treatment, such as diabetes care, oncology, cardiology, and radiology to name a few^10^.

DL has been successfully applied for several routine nephrology-related evaluation procedures^11–13^, which promises to improve speed and precision of pathologic workup of renal patients. Averitt et al. used CNNs to predict kidney survival, kidney disease stage, and various kidney function measures such as eGFR percentages from histological images^12^. Marsh et al. similarly used CNNs to predict glomerulosclerosis rates, i.e. the percent of glomeruli that are normal and sclerotic from frozen kidney transplant slides^13^. Both models were able to achieve performance on par with renal pathologists, thus demonstrating potential of DL systems to serve as a pathologist assistant. Yet there are many other kidney pathologies where no automatic quantification procedures exist, but would undoubtedly be of great benefit to patients, clinicians, and the entire health system.

In kidney allografts, inflammation is the defining feature of acute cellular rejection with various patterns of immune cell infiltrates. Lymphoid aggregates (LAs) are characterized by the recruitment of T, B, and dendritic cells and are visible in histological sections as a collection of distinct large cells with irregular nuclei and dark-blue cytoplasm^14^. LAs are commonly observed in patients with kidney allografts and their presence and localization may correlate with severity of acute rejection^15,16^. Patchy interstitial mononuclear cell infiltrates may be indicative of a milder form of alloimmune injury^17^. Mononuclear cell aggregates localized to the sub-capsular area or fibrotic tissue are usually interpreted as “non-specific” inflammation^17^. In addition to cellular rejection, various types of inflammatory infiltrates can also be seen in acute pyelonephritis, virus infections, and drug-induced interstitial nephritis^17^. Assessment of LAs is a time-consuming process with no accepted quantification standard. Despite a number of recent efforts in the domain of automatic quantification of renal features^11–13^, automated detection and quantification of inflammatory marks such as LAs in the kidney has not been previously attempted. Integration of digital pathology DL algorithms is especially crucial in situations where expert personnel are limited for expedient diagnosis such as in the setting of the interpretation of transplant kidney biopsies. Therefore, accurate automated diagnosis or flagging of inflammatory features could prove to be a valuable tool for pathologists to assess the underlying causes of renal dysfunction both in native and transplant kidneys, and has potential to improve precision and efficiency of renal biopsy analysis. In this study, for the first time, we present an efficient, accurate, and automated method of localizing LAs and measuring their densities in whole slide digital images of transplant kidney biopsies using convolutional neural networks. By helping to improve diagnostic accuracy and speed, we envision our method to be of great benefit to physicians supervising kidney transplantations.

## Methods

### Pathology material

A sample of transplant kidney biopsies collected between years of 2010 and 2016 at UCSF Department of Pathology from patients (*n*=61) presenting for evaluation of renal allograft dysfunction were analyzed. Hematoxylin and eosin-stained slides of the biopsies were digitized at 40x magnification with a Leica Aperio CS2 whole slide scanner, with final resolution of approx. 3960 pixels per mm, 15.68 µm^2^/pixel. The research using the retrospective data was approved by IRB #17-22317.

### Data Preparation

The 61 cases were split into a training set (*n*=44), a validation set (*n*=7), and a test set (*n*=10). The ground-truth labels for the LAs in five of the 10 test slides were provided by a board-certified renal pathologist. The other five test slides contained no LAs. The ground-truth labels for the training set and validation set were provided by an appropriately trained medical student. The training set was used to train the neural network and update the model parameters. The validation set was not used to update the model parameters, but was used to evaluate the model after each training epoch and identify the best generalizing model. The test set was used to gauge the final performance and generalizability of the trained model on slides not evaluated during training. Average slide size was 4.102 ± 0.259 billion pixels. To increase training efficiency and due to computational limitations, 1024×1024 pixel patches were obtained by grid-sampling from each slide only where kidney tissue was present. The training set thus consisted of 7669 patches, the validation set 1112 patches, and the test set 2200 patches after the grid-sampling. In the test set, 1239 patches were derived from the five LA-free slides and 961 patches were from the five LA-containing slides. To further increase training efficiency, our model was trained on 256×256 patches by down-sampling the 1024×1024 patches by a factor of four. During the evaluation on the test set, we performed test-time data augmentation ensembling to improve performance by averaging prediction over original and three flipped versions of images.

**Table 1.**
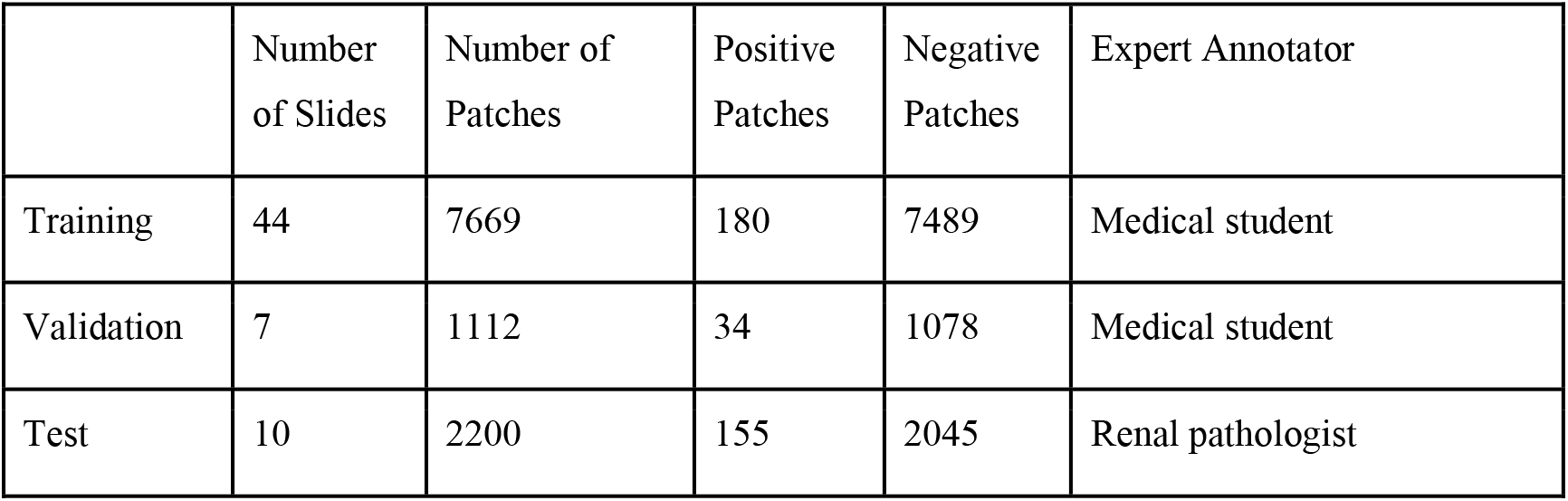
Number of biopsy slides, number of grid-sampled patches, number of patches with LAs, number of patches without any LAs, and qualification of the ground-truth label generator for the training, validation, and test sets.

### Model Architecture and Training

The slides were read and pre-processed using Openslide^18^ and Slideslicer^19^ python packages. For semantic segmentation, we used a modified version of U-Net ^6,20^, which consists of a sequence of contracting convolutional layers followed by a sequence of expanding convolutional layers. This allows for the final output to have the same resolution as the input while having a reduced number of parameters compared to an architecture without contracting and expanding paths. Unlike other encoder-decoder-like architectures, U-Net also combines feature maps from the contracting layers to the inputs for the corresponding expanding layers in a symmetric fashion. Our implementation used a VGG16 ^2^ head (including 13 convolutional layers with respective max-pooling layers) pre-trained on ImageNet dataset^21^ and a custom decoder, similar to model described in^20^ as shown in Fig. S5. The network was trained for 40 epochs, with each training epoch consisting of 7,669 iterations. The model was trained using a binary cross-entropy loss and the Adam optimizer with learning rate 10^−5^ and default parameters otherwise. Implementation is available on Github^22^.

### Model Evaluation

Based on the predicted segmentation probability map, a density score for each patch was calculated by counting the number of pixels where probability of LAs is higher than probability of normal tissue and dividing it by the total number of pixels in a patch. The intersection over union (IoU), Dice score, and area under the ROC curve were calculating using standard formulas described elsewhere^6,8,20^. As these metrics are either undefined or not meaningful when no true positive areas are present, they were first evaluated on LA-containing slides only, and mean and standard error across slides was reported. Next, these metrics were evaluated on the results from all slides combined together. The compactness of annotated and predicted regions of interest (ROIs) was evaluated using Polsby-Popper index (PPI) ^23^, with formula:

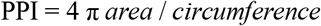

Unpaired predicted and annotated ROIs were compared using Mann-Whitney U test, and paired (true positive predicted and annotated) ROIs were compared using paired Wilcoxon signed rank test. The statistics are reported as mean ± standard error unless otherwise specified.

## Results

Using the trained model, we were able to accurately segment lymphoid aggregate regions, achieving an average area under the ROC curve of 97.78 ± 0.93%, an IoU score of 69.72 ± 6.24% and Dice score of 81.47 ± 4.69% across LA-containing slides in the test set. When aggregated predictions from both LA-containing and LA-free slides were considered, overall area under the ROC curve reaches 98.21%, IoU score 72.62%, and Dice score 84.14%. High-level inspection of predicted and observed LAs in whole slides in Fig. 1 reveals good overall agreement between predicted and observed patterns. This is further corroborated by high correlation between the predicted LA area fraction (2.40% ± 1.08% of total tissue area in test set slides) and the annotated LA area fraction (2.45% ± 0.98%) with Pearson R of 0.9756 (*p*=5.874e-8) as shown in Fig. 2. On a more granular level of image patches, annotated and predicted proportions of LAs per patch are also highly correlated (slide-level Pearson R = 0.9624 ± 0.0209 for LA-containing slides, Fig. S6).

**Figure 1.**
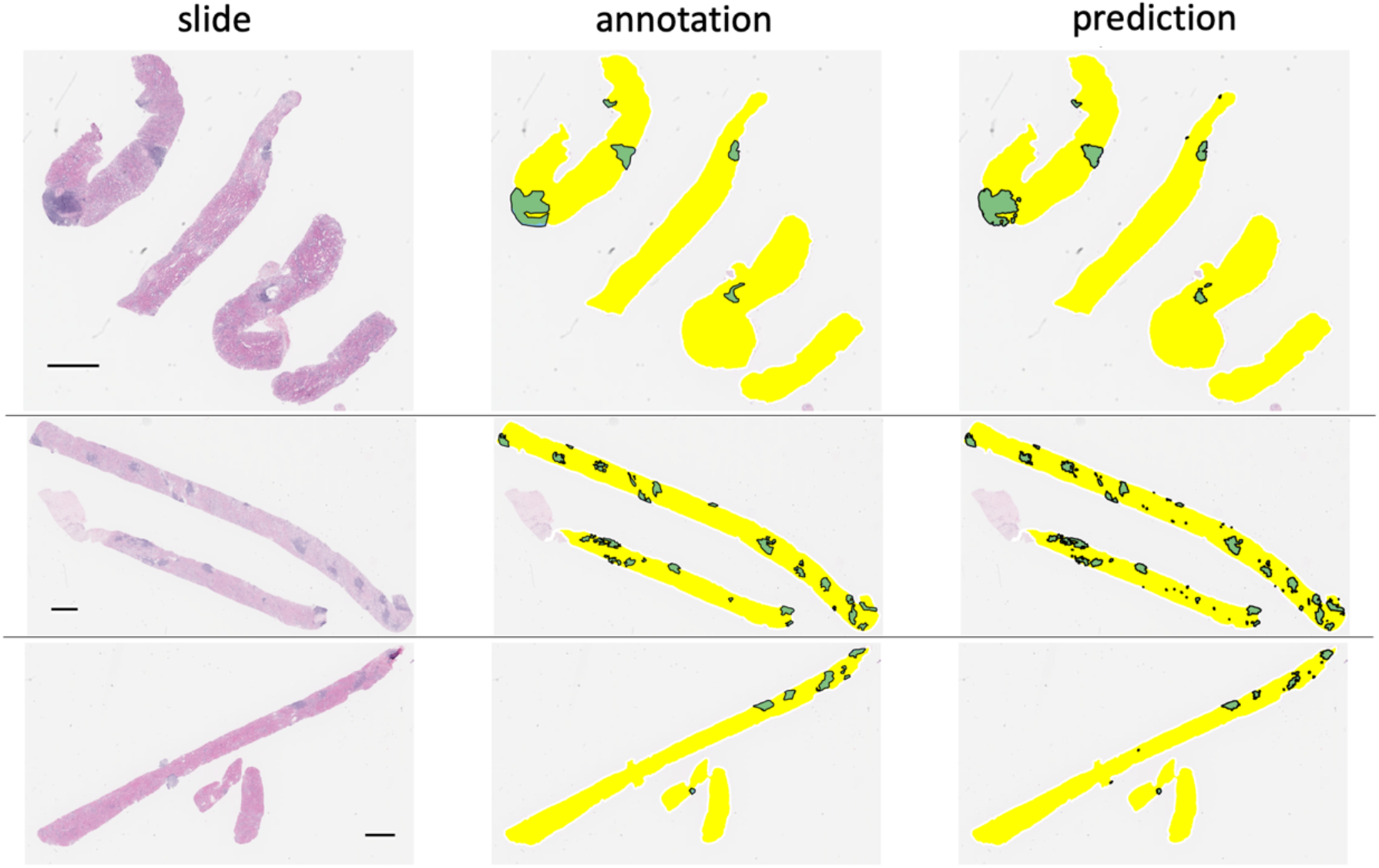
A coarse-level visualization of three representative core needle biopsy slides (left) alongside with LAs annotations provided by a renal pathologist (middle) and neural network prediction (right). Predictions with area less than 15,000 pixels are removed. The black horizontal bar indicates scale of 1mm.

**Figure 2.**
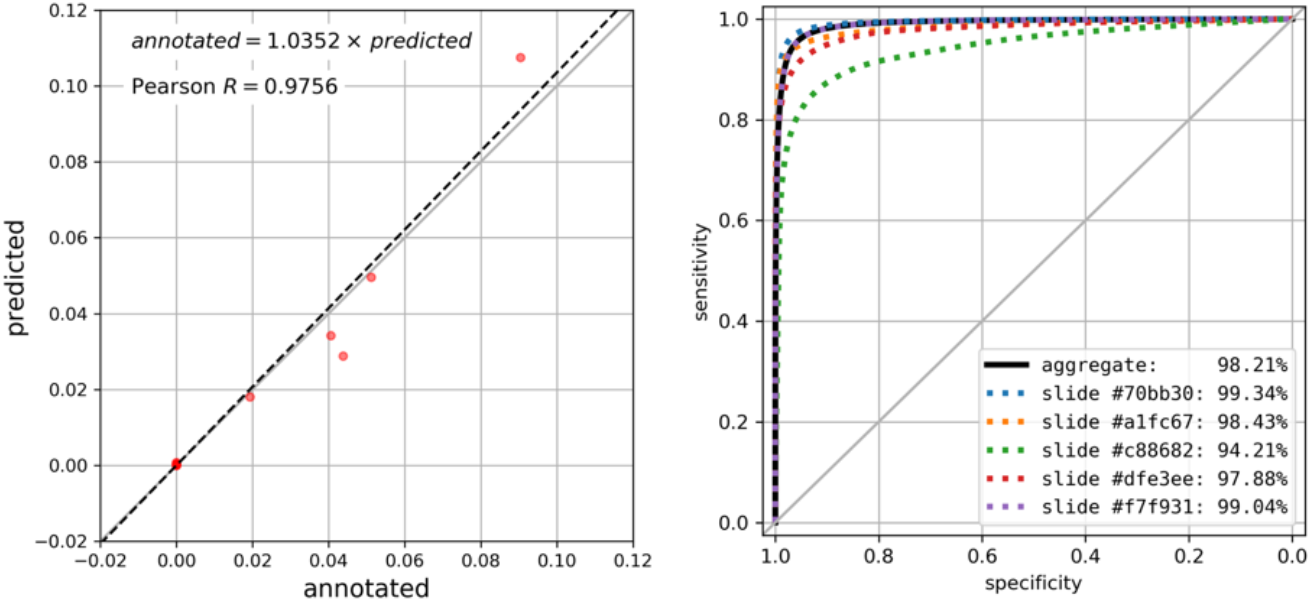
Performance metrics of the segmentation algorithm within the test set. **A**. Correlation plot of the area of LAs ROIs (as fraction of total tissue area) in annotations (*x*-axis) and predictions (*y*-axis), Pearson R of 0.9756 (*p*=1.5e-06). The regression line is shown in black dotted line, and regression equation is shown (*p*=5.874e-8). Note that 5 LA-free samples are densely clustered near the origin (0.0, 0.0). **B**. ROC curve for predicted probability of LAs-class pixels. ROC for all pixels aggregated across all samples is shown in black solid line (AUC=98.21, 95% confidence interval: 98.20% – 98.22% using DeLong method), and ROC for 5 individual LA-containing slides are shown in colored dotted lines. Diagonal grey line indicates ROC for random predictions.

Predicted LA areas were of wider range of sizes than annotated ones. Particularly, many small LAs were predicted compared to annotated ones. In order to further analyze the distribution of sizes and shapes of annotated and predicted LAs, we calculated areas and Polsby-Popper indices (PPI) characterizing the shape irregularity on the range from above 0 (highly irregular) to 1 (circular). The visualization of obtained metrics (Fig. 3) reveals presence of multiple small regular-shaped false-positive LAs predictions. An average area of the annotated LAs was 1,009,140 ± 134,419 pixels (64,339 ± 857µm^2^), while for predicted ones 301,600 ± 51,621 pixels (19,229 ± 3,291 µm^2^) in the test set. On the other hand, when only true positive predictions were matched with the annotations, the true positive areas were slightly larger than respective annotations, with regression equation: *predicted area* = 1.033 × *annotated area* (*p*<2.2e-16, Fig. 3B). The shape irregularity was similar in both groups on average (PPI = 0.4397 ± 0.0225 for annotated and 0.4418 ± 0.0098 for predicted, *p*=0.5376 in unpaired test). However, when only true positive predicted LAs were evaluated and paired with the annotations, the predicted LA regions have significantly less regular shape (PPI = 0.3165 ± 0.0180, *p*=2.557e-6 in paired Wilcoxon test), with regression equation: *predicted PPI* = 0.6707 × *annotated PPI* (*p*<2.2e-16, Fig. 3C).

**Figure 3.**
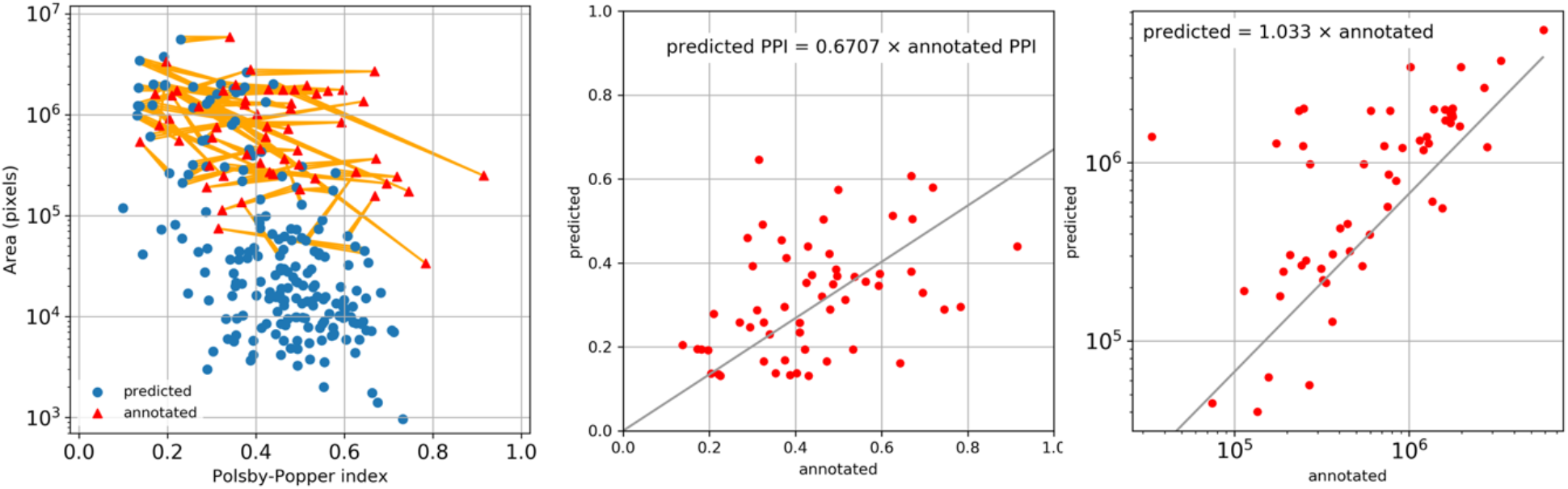
Comparison of size and shapes of annotated and predicted ROIs. **A**. Distribution of area and Polsby-Popper index (PPI) for annotation ROIs (red triangles) and prediction ROIs (blue dots). True positive predicted ROIs are connected to respective annotation ROIs with yellow lines. Note that false negative ROIs (with no connecting lines) mostly have small area. **B**. Correlation between the PPI of predicted and annotated ROIs (Pearson R=0.4184, *p*=0.0012). **C**. Correlation between the area of predicted and annotated ROIs (Pearson R=0.7961, *p*=1.3e-13).

As the minimal shape of annotated LAs was 33,782 pixels (2,153 µm^2^), with 5% percentile at 131,433 pixels (8,380 µm^2^), we sought to improve performance of the segmentation by removing small predicted LA areas. To this end, we calibrated IoU performance as a function of low-pass area threshold within the validation set (Fig. S7) and selected an optimal threshold for the area of predicted LAs that leads to maximal improvement in median IoU, at the value of 15,000 pixels (956 µm^2^). This thresholding improved the agreement between the counts of annotated and predicted LAs, thus reducing the mean absolute error from 13.0 to 6.57 LA regions per slide in validation set and from 14.1 to 6.7 LA regions per slide in the test set. At the same time, thresholding produced only a slight increase in IoU in LA-containing slides of the test set from 71.64% to 72.04%.

We show detailed visualization of correctly segmented, false negative, and false positive patches in Fig. 4A, 4B, and 4C respectively. In most cases, the segmentation outline produced by the model had high overlap with the human annotation (IoU = 69.72 ± 6.24%), but contained more spatial detail than the annotation. Oftentimes, smaller areas of fibrotic tissue or other non-lymphoid tissue that were included by the annotator into LA segmentation were excluded by the model, thus leading to false negative predictions (Fig. 4B). We saw that the model detected several smaller areas of lymphoid aggregates missed by the annotator initially (Fig. 4C, columns 1&2). Dense nuclear areas of small atrophic tubules or tangentially cut tubules that expose multiple nuclei in the same plane were sometimes mis-classified as LAs, which is especially common for predictions with small area (under 33,000 pixels; Fig. 4C, columns 3&4).

**Figure 4.**
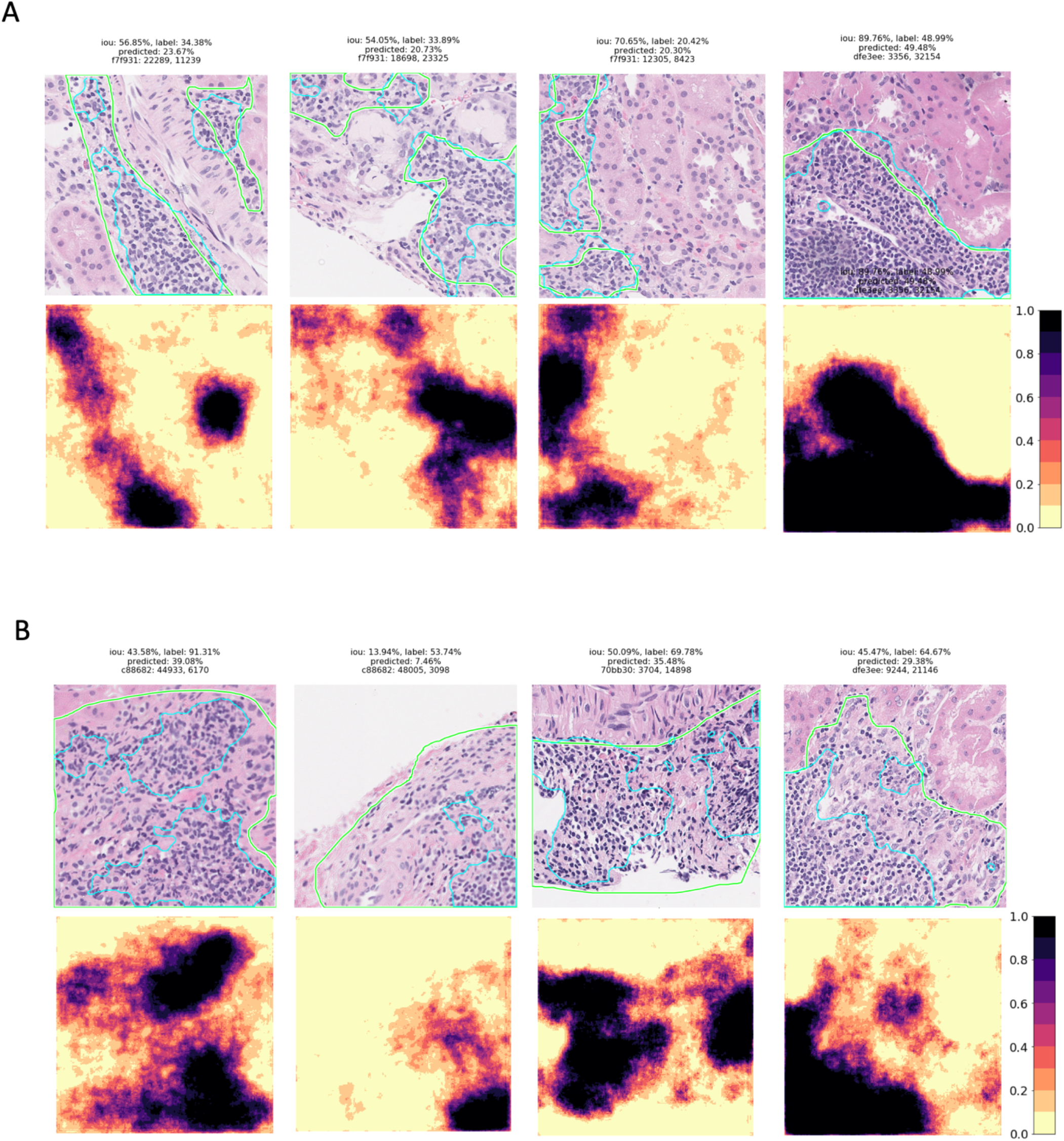
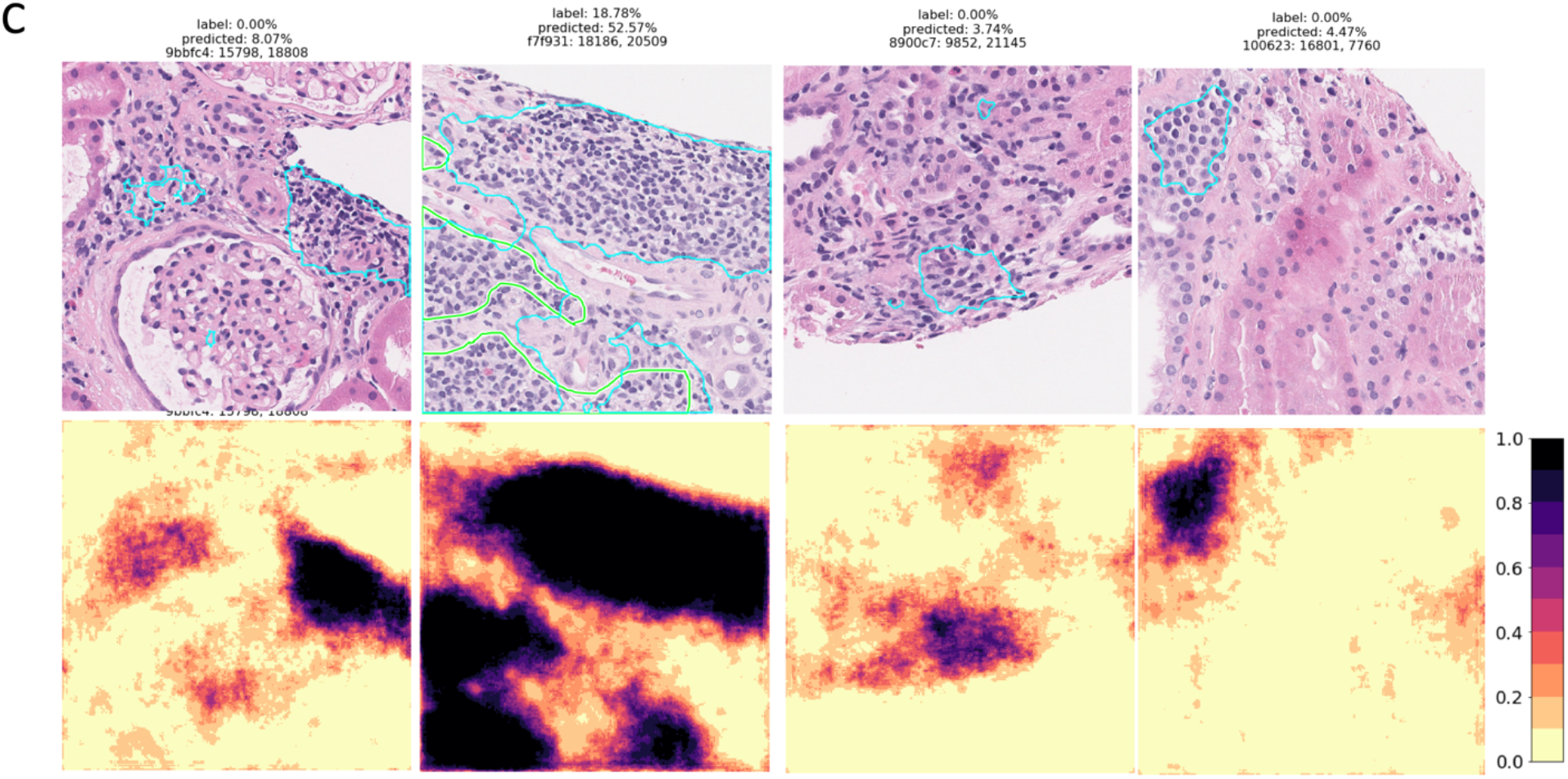
Segmentation of LAs in representative patches are shown with the original patch images and overlaid ground truth masks (green contour) and predictions (blue contour) in the top rows and segmentation probability heatmaps in the bottom rows. The IoU score, ground truth LA percentage, predicted LA percentage, slide ID, and patch coordinates are also displayed above each patch. **A**. Patches with good agreement between the annotation and prediction. **B**. Patches with false negative areas. **C**. Patches with false positive areas.

## Discussion

Our study is the first to demonstrate that a deep convolutional neural network can accurately identify lymphoid aggregates and provide a quantitative measure of inflammation in digitized histological slides. It shows that inflammatory markers can be efficiently and robustly quantified automatically using DL, and thus shows potential of DL algorithms in improving efficiency and precision of renal pathology workup. Our model produces annotations of a higher spatial detail than present in typical manual annotations, while generally agreeing with the pathologist’s annotations, as seen in Fig. 3A, and as indicated by a lower PPI index of true positive predicted LAs. Still the model produces numerous false positives of smaller size, that can be effectively removed by a low-pass area threshold filter. This shows that thorough expert-guided error analysis is necessary to keep medical DL algorithms unbiased, precise, and relevant to the problem at hand. On the other hand, our model detected areas of LAs initially missed by a pathologist (Fig.4C, first column). This showcases the robustness and the power of the neural network model compared to human annotator, given sufficient amount of training data. Additionally, the increased speed at which LAs can be detected with our method will free up time for the already encumbered clinician to focus on other necessary tasks of the procedure.

Visualization tools used in this work to display density of LAs, such as in Fig. 1, may be of potential use as a computer-aided decision support tool for pathologists and researchers investigating inflammatory processes in kidney allografts. As a next step, co-localization of lymphoid tissue with fibrotic and capsular tissue need to be learned as it is necessary for differential diagnosis of LA-associated conditions^17^. Furthermore, scope of future work will need to focus on assigning and predicting Banff scores for various types of kidney pathologies^24^. We believe our model, fine-tuned with respective labels, would be able to accurately predict Banff lesions and provide basis for score estimation once we have adequate ground truth labels. Such an algorithm, when implemented as a part of CAD system, could drastically speed up and simplify renal pathology analysis, as well as improve precision in clinics where specialized renal pathologists are not available.

## Data Availability

The code is available on Github and model weights are available upon request.

https://github.com/HadleyLab/kidney_lymphoid_aggregates

## Abbreviations

auROC: Area under the ROC curve
CAD: computer-aided diagnosis
CNN: Convolutional Neural Network
DL: Deep Learning
eGFR: estimated glomerular filtration rate
IoU: Intersection over the Union
LA: Lymphoid Aggregate
ROC: Receiver Operating Characteristic

## Acknowledgments

Research reported in this publication was supported by the National Institute of Health through the National Cancer Institute under award number UH2CA203792 and the National Library of Medicine under award number 1U01LM012675 (PI: DH). The content is solely the responsibility of the authors and does not necessarily represent the official views of the National Institutes of Health. AB is supported by a National Institute of General Medical Sciences training grant (5T32GM008440, PI: Judith Hellman)

---

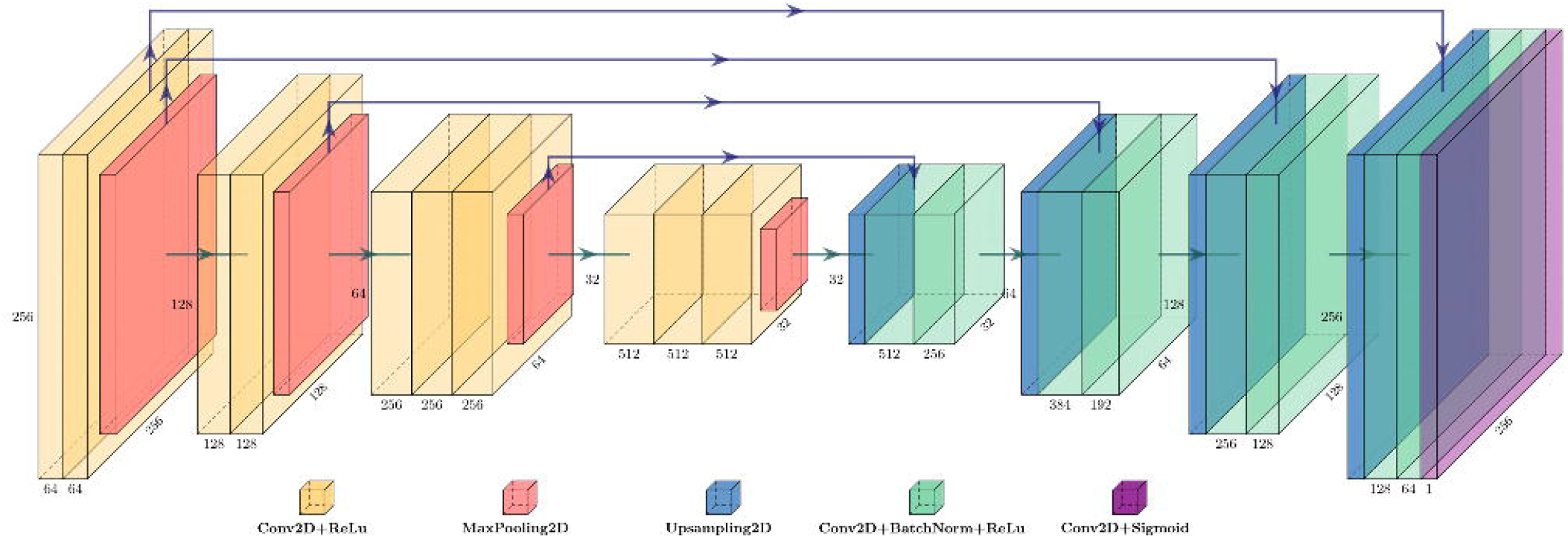

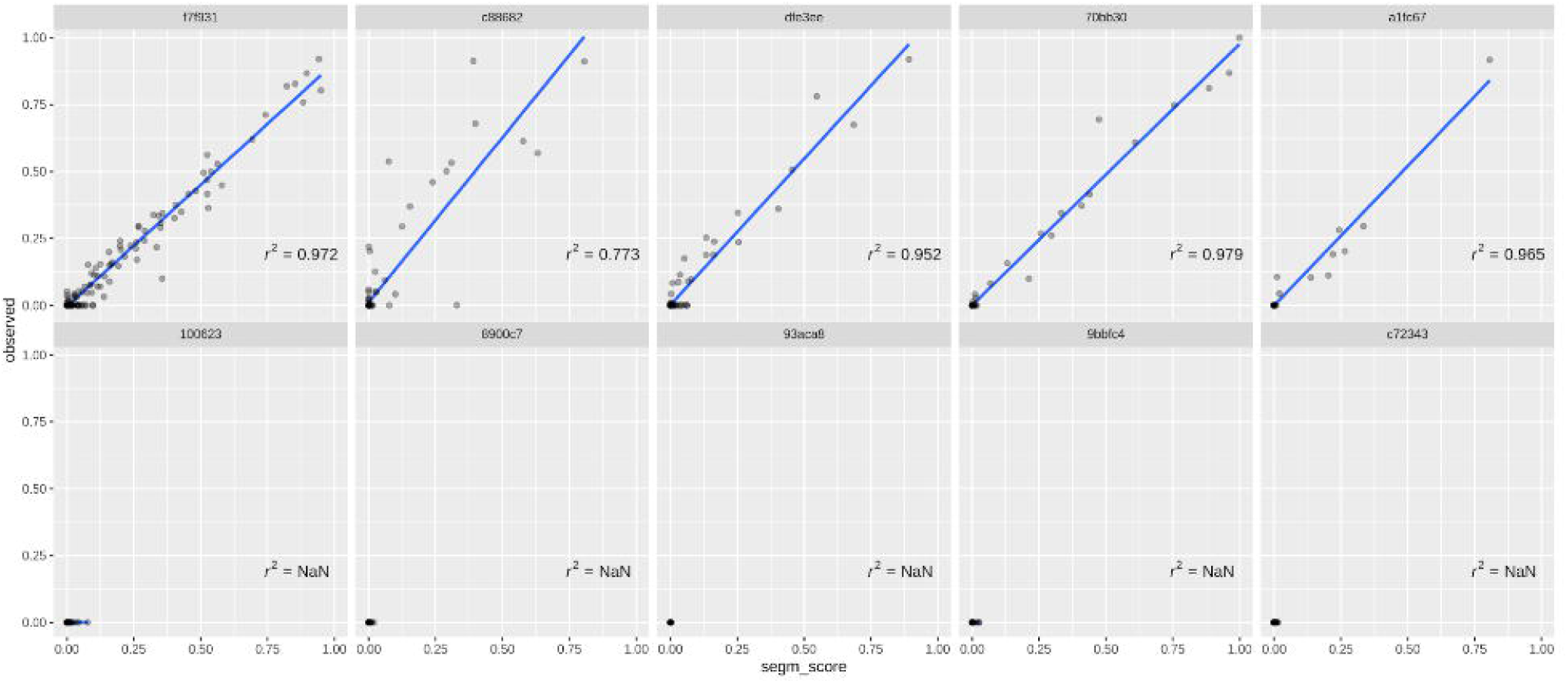

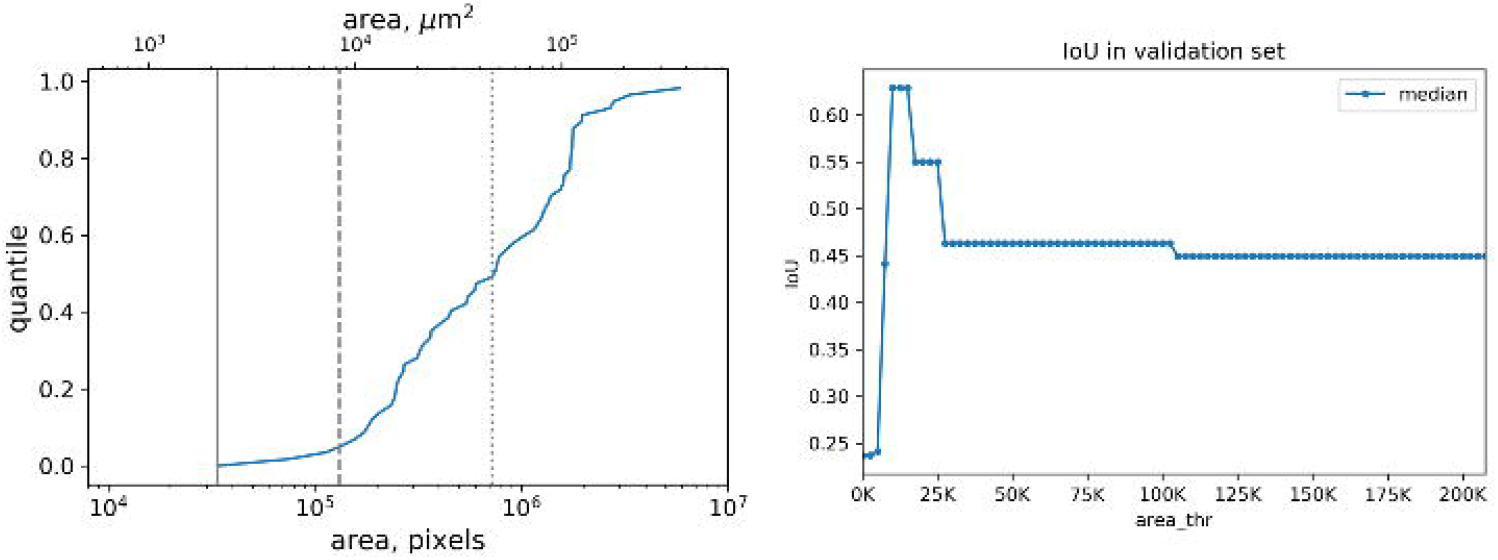

## Notes

### Competing Interest Statement

The authors have declared no competing interest.

### Author Declarations

All relevant ethical guidelines have been followed and any necessary IRB and/or ethics committee approvals have been obtained.

Any clinical trials involved have been registered with an ICMJE-approved registry such as ClinicalTrials.gov and the trial ID is included in the manuscript.

